# Ask your big brothers and sisters! A study protocol on multidisciplinary exploratory research into a preventive community youth empowerment program using biopsychosocial measures

**DOI:** 10.1101/2025.10.07.25337469

**Authors:** Marleen Verhagen, Carin Rots, Linda M.A. van Loon, Marjolein Stoopendaal, Lieke G.M. Raaijmakers, Anja C. Huizink, Sjoerd A.A. van den Berg, Jolanda J.P. Mathijssen

## Abstract

**Introduction:** Children growing up in adversity often experience setbacks and toxic chronic stress. Throughout the particularly vulnerable developmental phase of adolescence, chronic stress can lead to significant mental and physical repercussions. However, these adverse outcomes can also be prevented or alleviated.

There are indications that preventive programs with an integrated approach can contribute to reducing chronic stress. Big Brother, Big Sister (BBBS) is a preventive community Youth Empowerment Program implemented in low socio-economic status neighborhoods. Developed by and for young people, it empowers participants to reconnect with education and employment through professionally facilitated peer group activities. The program is embedded in local networks.

**Main objectives:** To assess the applicability of the following biopsychosocial outcome measures—stress(ful life events), subjective psychological well-being, and social participation—within a diverse group of young people in vulnerable positions engaged in the BBBS program. Additionally, to provide insight into how these outcomes change over time.

**Methods and Materials:** This exploratory quantitative study has an observational design with three measurements (at the onset of the program, after 6 months of participation and at outflow). Approximately 80 young people aged 14 to 27 will participate in this study.

Various methods will be utilized to assess outcomes; questionnaires will be completed (i.e. the Perceived Stress Scale; Basic Psychological Need Satisfaction and Frustration Scale, Stressful Life Events Scale, social participation and general background (hair) characteristics questionnaires), and hair samples will be collected to conduct cortisol and cortisone analyses, as proxy for prolonged physiological stress.

**Discussion:** The multidisciplinary collaboration of partners within the research group combined with the biopsychosocial approach of this study is innovative and provides opportunities for cross-domain learning and collaboration to pursue (research) opportunities for evidence-based (early life) stress prevention and improvement of psychological well-being during the critical developmental phase of adolescence.

## Introduction

### Early Life Stress (ELS) and resilience in adolescence

Children and young people growing up in adversity often experience setbacks and toxic chronic stress [1] often referred to as Early Life Stress [2,3]. Toxic chronic stress occurs when the stress system is repeatedly reactivated without sufficient opportunity for recovery. Throughout the particularly vulnerable developmental phase of adolescence (10-24 years) [4], persistently elevated levels of stress hormones can significantly affect (future) mental and physical health, learning, and behaviour through gene-environment interactions and can result in neurobiological changes [1,2,3,5,6]. Chronic stress also modifies the stress response; individuals react more quickly and violently to a stressor later in life, and the stress response persists longer [1,6]. As the brain undergoes rapid growth and development in adolescence, the adverse outcomes resulting from chronic stress can be prevented or alleviated during this period and transformed into more favorable development by enhancing well-being and fostering resilience [5,7]. This way, the plastic brain can be reprogrammed [8]. Since individuals who have experienced stress in early life often struggle to cope with stress induced by their own children [1,3], it is crucial for the (mental) health of the next generation that adverse outcomes resulting from toxic chronic stress in early life are prevented, alleviated, or transformed during adolescence.

### Hair cortisol, hair cortisone and chronic stress

As a human response to acute and chronic stress, the hypothalamic-pituitary-adrenal (HPA) axis is activated. When repeatedly activated under chronic stress, cortisol is released to an increased extent [9,10]. Nonetheless, personal characteristics such as resilience are associated with a reduced activation of the HPA axis [11] and “dysregulated” HPA axis functioning can result in either increased or decreased cortisol levels [12].

Measurements of both cortisol and cortisone (inactivated cortisol) in human hair are increasingly utilized as biomarkers to gain insight into chronic stress exposure. Hair steroid measurements provide retrospective insights because the long-term cumulative release of cortisol and cortisone can be measured up to several months before [13].

Hair cortisol concentration (HCC) has been associated with adult mental health in previous research; for example, increased cortisol levels have been found in adults experiencing ongoing chronic stress, while reduced hair cortisol levels have been found in adults with anxiety disorders such as PTSD [12,14]. Although this research in children and young people, especially for those in vulnerable positions, is in its early stages, a recent review and meta-analysis by Li et al. [15] demonstrated that chronic stress was positively correlated with HCC. Hence, hair cortisol could serve as a biomarker for chronic stress among children and young people (aged 5-21 years). Recently, reference ranges for hair cortisol in children (aged 0-18 years) have been identified [16]. This may provide an opportunity to compare the results of this study with these reference ranges.

In addition to cortisol, the hormone cortisone might be a useful biomarker for stress research. Very few studies in young people include measurements of both hair cortisol and cortisone [17]. Results from the study by Ullmann et al. [18] showed that the hair cortisol and cortisone concentrations were significantly correlated with mental and physical stress, as well as with stress perception in students (aged 18-35 years).

Research on the association between socio-economic status and hair cortisol and cortisone levels showed that a low neighborhood-level socio-economic status was significantly associated with higher hair cortisol and cortisone levels in children and adolescents (aged 4-18 years) [19].

This study will provide insight into experienced (early life) stress, subjective psychological well-being, social participation, and the hair cortisol and cortisone levels of a group of young people of mixed descent who primarily grew up in low SES neighborhoods, are facing difficulties in connecting with society, and participated in the Big Brother, Big Sister program.

### Community Youth Empowerment Program “Big Brother, Big Sister” Breda

‘Big Brother, Big Sister’ (BBBS) is an integrated community Youth Empowerment Program (YEP) that empowers participants to reconnect with society and make progress in fostering positive social connections and developing (psychosocial) skills to pursue meaningful goals [20]. The program is peer-developed by and for young people aged 14–27; Big Brothers and Sisters of diverse backgrounds and often with vocational education. Building trust between workers and participants is key, offering young people easy access to a safe space to reconnect with education or employment and grow personally and socially with peer support. Big Brothers and Sisters serve as role models, improving life in the neighborhoods where they are active and primarily grew up. BBBS supports around 150 youths annually across three vulnerable neighborhoods in Breda. Figure 1 provides further details about the program.

**Figure 1.**
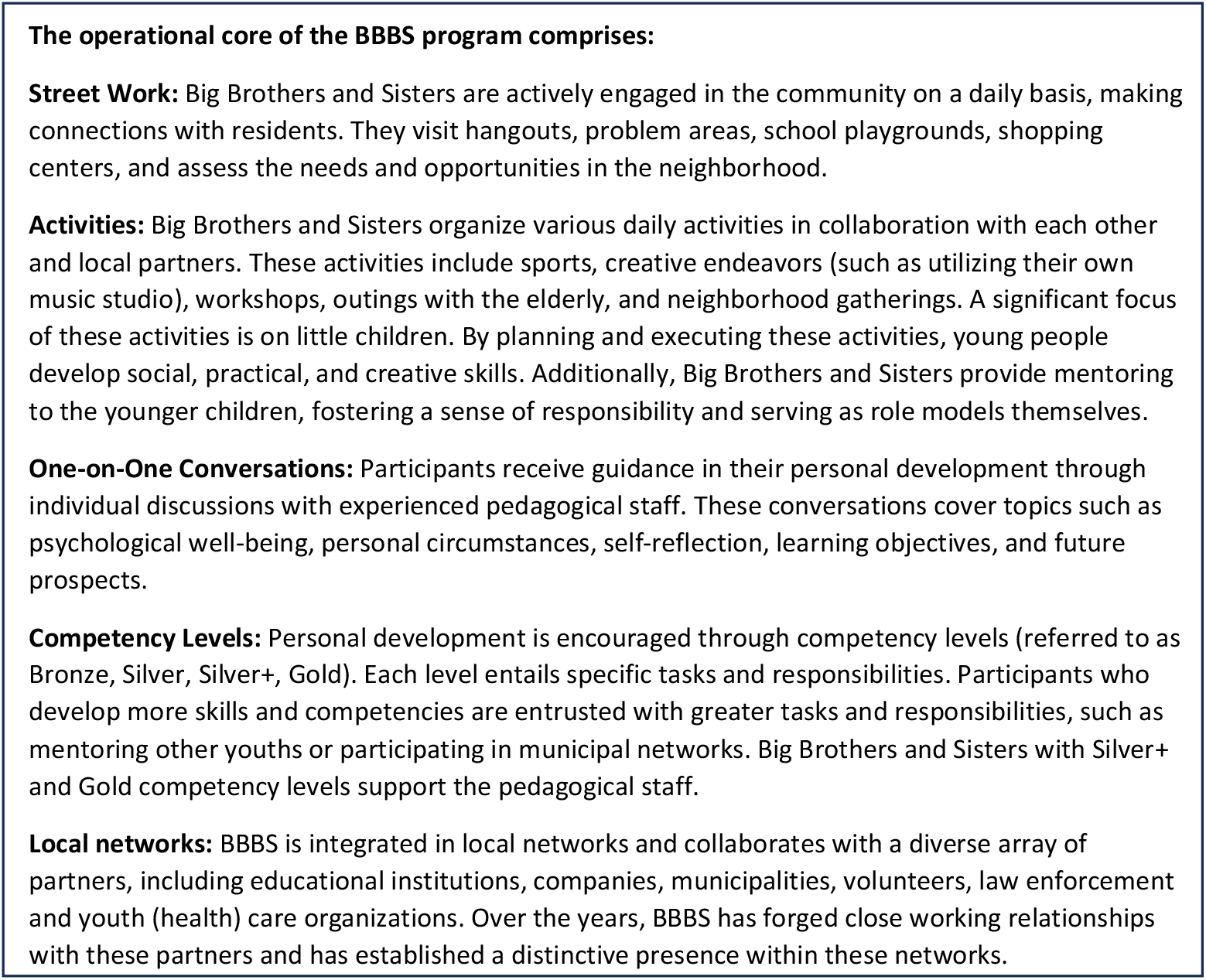
The operational core of the BBBS program.

### The operational core of the BBBS program comprises

#### Street Work

Big Brothers and Sisters are actively engaged in the community on a daily basis, making connections with residents. They visit hangouts, problem areas, school playgrounds, shopping centers, and assess the needs and opportunities in the neighborhood.

#### Activities

Big Brothers and Sisters organize various daily activities in collaboration with each other and local partners. These activities include sports, creative endeavors (such as utilizing their own music studio), workshops, outings with the elderly, and neighborhood gatherings. A significant focus of these activities is on little children. By planning and executing these activities, young people develop social, practical, and creative skills. Additionally, Big Brothers and Sisters provide mentoring to the younger children, fostering a sense of responsibility and serving as role models themselves.

#### One-on-One Conversations

Participants receive guidance in their personal development through individual discussions with experienced pedagogical staff. These conversations cover topics such as psychological well-being, personal circumstances, self-reflection, learning objectives, and future prospects.

#### Competency Levels

Personal development is encouraged through competency levels (referred to as Bronze, Silver, Silver+, Gold). Each level entails specific tasks and responsibilities. Participants who develop more skills and competencies are entrusted with greater tasks and responsibilities, such as mentoring other youths or participating in municipal networks. Big Brothers and Sisters with Silver+ and Gold competency levels support the pedagogical staff.

#### Local networks

BBBS is integrated in local networks and collaborates with a diverse array of partners, including educational institutions, companies, municipalities, volunteers, law enforcement and youth (health) care organizations. Over the years, BBBS has forged close working relationships with these partners and has established a distinctive presence within these networks.

### In- and outflow at BBBS program

Young people enter the program through various channels, including direct contact with Big Brothers and Sisters, referrals from community partners such as welfare organizations, housing associations, and youth health care services, as well as through schools. Additionally, young people can register themselves for the program. An interview is conducted with each young person to assess their eligibility for participation in which factors such as their willingness to develop, learn, and serve as a role model (as a ‘Big Brother’ or ‘Big Sister’) are considered essential for inclusion. When participants achieve their goals, find employment or make progress in school, they typically exit the BBBS program. Participants remain engaged in BBBS for a period ranging from six months to two years.

### Previous research and effective elements/ theoretical underpinning

The BBBS program has been implemented for 15 years. Throughout its development, BBBS has undergone initial internal and external evaluations [21], which have been used to refine the program.In 2019, a qualitative study on Big Brothers’ self-perceived effects of the program revealed that participants feel comfortable and respected within BBBS and reported progress in fostering positive social connections and developing (psychosocial) skills to pursue meaningful goals [20].

Achievements include reduced aggression, improved cooperation, enhanced sense of responsibility, and a sense of doing something meaningful for others and for themselves. The way BBBS is designed aligns well with Self-Determination Theory. Self-Determination Theory posits that psychological well-being is determined by three basic human needs: autonomy, positive connection with others, and competence to achieve meaningful goals [22]. There are indications that preventive programs with an integrated approach, meeting these basic human needs, can increase well-being and contribute to reducing chronic stress [3,8]. The following effective elements appointed by Pijpers et al. [3] are integral to the preventive Youth Empowerment Program BBBS:

1. Eliminating harmful stressors (e.g., aggressive encounters in the street, exposure to criminal pressures); 2. Enhancing (psychosocial) skills, autonomy and competencies, thereby boosting personal resilience and enhancing future opportunities; 3. Providing a social environment characterized by positive social interactions and connections with the community. Within a secure pedagogical climate, young people can confront challenges, fostering confidence in themselves and others.

### Main objectives

Overall, ‘Big Brother, Big Sister’ serves as a preventive community Youth Empowerment Program with the potential to mitigate early-life stress while enhancing psychological well-being and social engagement. Therefore, BBBS emerges as a promising environment to assess the applicability of the following biopsychosocial outcome measures—stress(ful life events), subjective psychological well-being, and social participation—within a diverse target group of young people in vulnerable positions engaged in the BBBS program and to provide insight in how these outcomes change over time.To our knowledge, there has been hardly any multidisciplinary research on preventive youth programs in low socioeconomic status (SES) neighborhoods, such as BBBS, using biopsychosocial outcome measures that include (early life) stress, psychological well-being and social participation of youth both at the beginning of a program and over time. Recent literature underscores the particular necessity for biopsychosocial research among young people in vulnerable positions and of mixed descent. Such research is crucial for elucidating stress mechanisms and for pursuing evidence-based strategies aimed at preventing early-life stress and enhancing psychological well-being during the sensitive developmental period of adolescence [1,3,6,16].

## Methods and materials

### Design

The quantitative study is exploratory in nature, employing an observational design with three measurement points: at the onset of the program, after 6 months of participation, and at outflow. Recruitment commenced on November 15^th^, 2021, and is set to conclude on May 31^th^, 2025.

### Research population - involvement and recruitment

All young people who enrolled in the program between the end of October 2021 and December 2023 were invited to participate in the study. Preparatory discussions at BBBS, estimated participation of 80 young individuals aged 14-27. Written informed consent is required at each measurement, with parental consent for those aged 14-16. Once consent is obtained, participants will be invited to measurement sessions at their respective BBBS location, situated across three neighbourhoods in Breda. These sessions will be conducted in private settings and scheduled at mutually convenient times. Participants will receive compensation in the form of shopping vouchers: €10 for the first two sessions and €20 for the third, as determined in consultation with BBBS.

### Inclusion criteria

The study will include all young people meeting the following criteria:

- Enrolled in BBBS between the end of October 2021 and December 2023.
- Aged between ≥ 14 years and ≤27 years.
- Demonstrate willingness to develop, learn, and serve as a role model within the program (as a ‘Big Brother’ or ‘Big Sister’).

### Exclusion criteria

The study will exclude all young people who have participated in BBBS < 1 years ago (determined at baseline).

### Procedures

Participants will be invited for measurement within four weeks after the start of the program, after 6 months, and at outflow or when they achieve their original starting goals (sometimes they continue in the program in another position). The measurement process takes approximately 20 minutes and includes self-reported questionnaires covering perceived stress, stressful life events, and psychological well-being. Questionnaires regarding social participation, general background and hair characteristics, will be verbally administered by the researcher.

With permission, a small strand of approximately 100 hairs is discreetly cut with nail scissors from the posterior vertex of the scalp, ensuring minimal visibility by selecting hair from a deep layer. These hair samples will be analysed for cortisol and cortisone levels [13]. Table 1 provides further details about the timing of the various measurement moments.

**Table 1.**
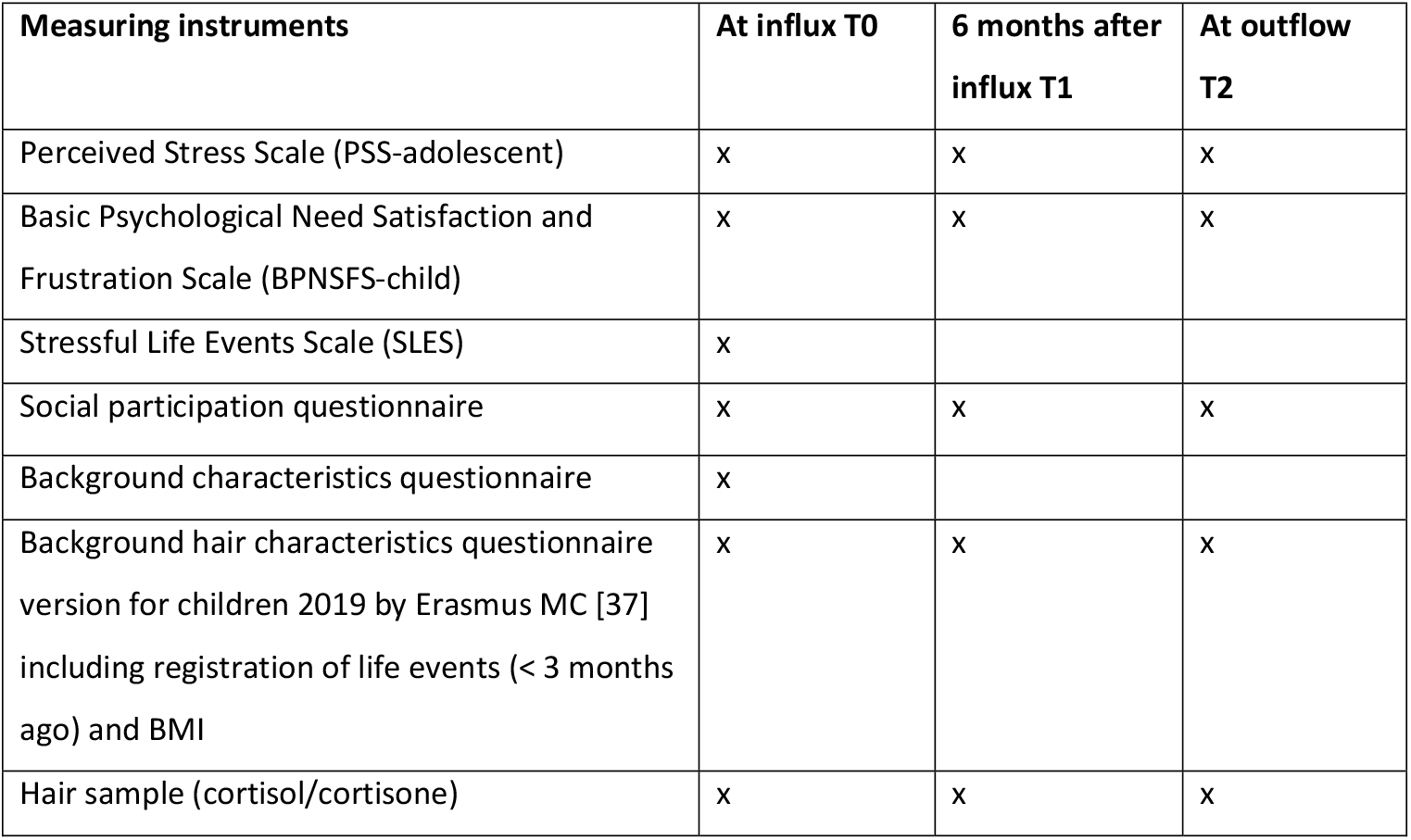
Measuring instruments per measuring moment.

| Measuring instruments | At influx T0 | 6 months after influx T1 | At outflow T2 |
| --- | --- | --- | --- |
| Perceived Stress Scale (PSS-adolescent) | x | x | x |
| Basic Psychological Need Satisfaction and Frustration Scale (BPNSFS-child) | x | x | x |
| Stressful Life Events Scale (SLES) | x |  |  |
| Social participation questionnaire | x | x | x |
| Background characteristics questionnaire | x |  |  |
| Background hair characteristics questionnaire version for children 2019 by Erasmus MC [37] including registration of life events (< 3 months ago) and BMI | x | x | x |
| Hair sample (cortisol/cortisone) | x | x | x |

### Ethical considerations and approval

All data in this study will be kept confidential and pseudonymised, with no names or personal information linked to the answers or hair measurements.

Participants may skip certain questions or withdraw from the study at any time, without providing a reason or facing any consequences. A pedagogical supervisor is available on-site to support the participant if needed. This study was approved by the Medical Ethics Review Committee Brabant on November 1, 2021, and was found not to be subject to the Medical Research Involving Human Subjects Act (WMO) (METC nr: NW2021-91).

### Measures

#### Perceived stress

The ten-item Perceived Stress Scale (PSS-adolescent) [23] will be utilized to measure the extent to which respondents experience stress due to unpredictability, lack of control, and overload (e.g., How often have you felt stressed and nervous? Rated on a five point Likert scale). A Dutch version for adolescents is available. Previous studies have demonstrated a Cronbach’s alpha > .70 [24,25].

#### Subjective psychological well-being

The twenty-four-item Basic Psychological Need Satisfaction and Frustration Scale (BPNSFS-child) [26], utilizing a five point Likert scale, will be employed to subjectively measure the satisfaction and frustration of basic psychological needs in one’s life (e.g., I often doubt whether I am good at things). This scale is grounded in the Self-Determination Theory [22]. The Dutch version of BPNSFS-child is comprehensible for a diverse range of young individuals, as confirmed by a pretest involving approximately fifteen Big Brothers and Sisters (28-06-2021). In a sample of 359 Belgian adolescents (mean age 17.87), Cronbach’s alphas for autonomy, relatedness, and competence satisfaction were 0.69, 0.77, and 0.81 respectively [26].

#### Stressful Life Events

A questionnaire based on the Stressful Life Events Scale (SLES) [24,27] will be utilized, comprising twenty-six events ranked by the degree to which the event aggravates the lives of children (e.g.,divorce of parents or being a victim of violence). Participants will indicate whether or not they have experienced each event during their lifetime.

#### Social participation

Social participation is operationalized as involvement in education, employment, volunteer work, membership in (sports) clubs, physical activity, gaming, and conflicts or fights (yes or no and frequency in the past 3 months). A questionnaire, developed with BBBS, measures social participation using relevant questions from existing youth monitors conducted by the Municipal Health Service West-Brabant. Additionally, the questionnaire assesses competence levels within BBBS (see Figure 1).

#### Background characteristics

The background characteristics questionnaire includes items such as gender, age, type of education, grade, completed education, place of residence, neighborhood where one grew up, ethnicity, country of birth (of father and mother), living situation, financial situation, and a number of questions about participation in BBBS, which will be administered verbally.

#### Background hair characteristics

The background questionnaire on hair characteristics, based on the 2019 version for children from Erasmus MC, will include questions about hair color, dyeing/blonding/permanent treatments, frequency of washing, degree of excessive sweating on the head, use of hair products, use of corticosteroids, use of contraception, drug use, alcohol use, and general registration of notable life events in the past 3 months (“Have you experienced any stressful life events in the past 3 months?”). Additionally, length and weight will be inquired. This questionnaire will be administered verbally and is a required part of the application for hair cortisol/cortisone analysis. Questions regarding these background characteristics will only be asked when the hair analysis is conducted, except for the general registration of stressful life events in the past 3 months, which will be asked of all participants regardless of their participation in the hair analysis.

#### Hair sample

The hair analysis will be conducted using an improved LC-MS/MS-based method [13]. Hair grows at an average rate of 1 cm per month. Therefore, the 3 cm hair sample closest to the scalp measures the cumulative cortisol/cortisone (chronic stress level) over the past three months prior to each measurement [28].

An overview of the measuring instruments per measurement moment can be found in Table 1. At T0, all instruments will be administered. At T1 and T2, the registration of general background characteristics and Stressful Life Events will not be measured again.

### Data analysis

First, background characteristics, experienced (early life) stress, subjective psychological well-being, social participation, and the hair cortisol and cortisone levels of the respondents at T0 will be described and, if possible, compared to average Dutch young people. Chi-squared analyses, independent sample t-tests, and ANOVA will be used accordingly to assess differences in outcome variables for background variables (such as ethnicity, age, gender) and specific hair characteristics. Multivariable regression analyses will be performed to assess associations between background characteristics and outcome measures at T0.

Linear mixed models are then used to measure changes over time in experienced (early life) stress, subjective psychological well-being, social participation, and the hair cortisol and cortisone levels of the respondents individually and relative to each other and to determine whether these changes are related to background characteristics.

### Data management

The personal data of the participant will be encrypted and stored under a randomly assigned respondent number. Completed questionnaires will then be pseudonymized. Only the principal investigator will have access to the translation key to decrypt the questionnaire. The questionnaires collected as part of the study will be digitized into the Enalyzer program and stored separately in sealed envelopes in a locked room. Hair samples will be kept in sealed envelopes marked with a respondent number in a locked room and brought to the laboratory by the researcher in batches.The data will be used for scientific purposes only and will be destroyed 10 years after the completion of the study (2035). Data management agreements have been made with collaborating partners and the laboratory, which are documented in an overall consortium collaboration agreement and a specific collaboration agreement with the laboratory concerning the analyses of cortisol and cortisone. These collaboration agreements and METC-assessment have been reviewed by the privacy officer and data protection officer of the Municipal Health Service GGD West-Brabant.

## Discussion

The Big Brother, Big Sister (BBBS) program is a promising preventive youth empowerment initiative targeting mixed-descent adolescents from low socio-economic neighborhoods who face societal integration challenges. This group is typically underrepresented in research, particularly in studies concerning adolescent mental health and the measurement of hair cortisol and cortisone [16,29]. Due to its proven practical value over 15 years, further scientific substantiation is desired.

Conducting multidisciplinary research within the BBBS setting presents a valuable opportunity to deepen our understanding of stress mechanisms and the utilization of biopsychosocial outcome measures among a diverse group of young individuals in vulnerable circumstances. The selected approach for this quantitative study is an observational longitudinal design with an exploratory focus, integrating biopsychosocial measures. This methodology aims to provide insights into the relevance and feasibility of impact studies for future research. Starting with an exploratory perspective, such a phased approach is recommended for studying the outcomes of integrated, practice-oriented interventions [30].

Research within this practical program highlights challenges, such as small participant numbers. Secondly, adolescents that will be included live in a dynamic and multifactorial context which is beyond the researchers control. Additionally, participants facing challenges with societal integration may be more vulnerable to dropping out, which could complicate the completion of all three measurements. Finally, there is a temporal mismatch between hair cortisol/cortisone levels, which reflect an average over the past three months, and questionnaire outcomes, which capture the current status or the past four weeks.

Nonetheless, the study’s strengths lie in its close collaboration with both professional workers and a diverse group of young participants with lived experiences. This practical knowledge is complemented by a multidisciplinary scientific network, which combined with the biopsychosocial method, represents an innovative approach. This integrated approach fosters cross-domain learning and collaboration, paving the way for evidence-based strategies to mitigate early-life stress and enhance adolescent well-being.

## Data Availability

The data of this study are not publicly available due to privacy and ethical restrictions.

## Acknowledgements

The authors would like to acknowledge the professional workers of ‘Grote Broer, Grote Zus Breda’ for their contribution to the study.

## Supporting Information

not applicable.

Authors declare no competing interests.

## Funding

This work was supported by Stichting Delen geeft meer, project number 2021/11/01; https://delengeeftmeer.nl/ The research design was finalized prior to receiving funding from the foundation. One of the authors, C. Rots, was involved in the research design, while employed at the municipal health service of West-Brabant. Currently she is working at Stichting Delen geeft meer. In this role she solely contributed to the revision of this study protocol.

The data of this study are not publicly available due to privacy and ethical restrictions.

